# Accuracy and Correlation Between Bed Scale and Standing Scale Weights in Monitoring Volume Status in Patients Hospitalized for Acute Decompensated Heart Failure

**DOI:** 10.1101/2023.08.25.23294652

**Authors:** Devin Skoll, Phillip Abarca, Vu Pham, Anushka Das, Clark Mantini, Han Tun, Helga Van Herle, Ajay Vaidya, Aaron M. Wolfson, Michael W. Fong

## Abstract

**Background:** Accurate monitoring of volume status is vital in managing diuretic therapy in patients with acute decompensated heart failure (ADHF). Guidelines recommend daily measurement of standing body weight as an indicator of volume status, though many healthcare facilities use bed scales instead. There is limited data surrounding the accuracy of bed scales compared to standing scales, and literature suggests discrepancy between the two. This study is the first to evaluate the accuracy and correlation between scales among patients hospitalized for ADHF.

**Methods:** Adults (age ≥18) diagnosed with ADHF and its derivatives who were hospitalized at our center between March and April 2023 were identified via electronic medical record. Patients were included if they demonstrated capacity to consent and were medically cleared to ambulate independently. Participants’ weights were measured via bed scale and standing scale at the same time on each day of data collection. The mean difference (bias), precision [standard deviation (SD)], and limits of agreement (±2 SD) were calculated and graphed using the Bland-Altman method. Based on expert opinion, a bias of greater than ± 0.6 kg and precision greater than ± 3.0 kg were considered clinically significant.

**Results:** 51 pairs of bed and standing scale weights were obtained among 43 patients. The mean difference ± precision between weights was 1.42 ± 1.18 kg (95% CI, -0.894 – 3.73), exceeding the recommended bias. In addition, 71% (n=36) of participants’ weights differed by more than 0.6 kg between the bed and standing scale, 22% (n=11) differed by more than 2.0 kg, and 5.9% (n=3) differed by more than 4.0 kg (see Table I).

**Conclusions:** Bed scales are not accurate compared to standing scales and may lead to errors in fluid management in patients hospitalized for acute decompensated heart failure.

**What is Known:**

- Adequate diuresis and volume status optimization are mainstays of therapy in patients with acute decompensated heart failure
- Guidelines recommend measurement of standing body weight as an indicator of volume status, though many hospitals use bed scales instead

**What the Study Adds:**

- This is the first study to assess the difference in weights between bed and standing scales in patients hospitalized for acute decompensated heart failure
- This study provides evidence that bed scale weights are not accurate compared to standing scale weights in this population

## Introduction

Heart failure (HF) is a common medical condition affecting nearly 6.5 million Americans.^1^ Acute decompensated heart failure (ADHF) accounts for over 1 million inpatient admissions in the United States every year and results in almost $40 billion in healthcare costs.^2^ Unfortunately, ADHF is the most common cause of Medicare readmissions with a 30-day readmission rate of roughly 23%.^3^ This not only impacts hospital reimbursement rates, but also increases overall costs to the healthcare system. Adequate diuresis and volume status optimization are mainstays of therapy and achieving optimal volume status prior to discharge is critical in lowering readmission rates and is associated with improved mortality. Accurate in-hospital weight assessment is vital in determining volume status. Current AHA guidelines recommend measurement of standing body weight at the same time each day during hospitalization to monitor HF treatment.^4^ However, bed scales are commonly used in hospitals instead, as standing scales require patients to be ambulatory. This is often not possible when patients are bedbound, sedated, deemed a high fall risk, or receiving medical therapy that hinders ambulation (i.e. dialysis, intra-aortic balloon pumps, Purewick urinary catheters). Limited data exist regarding the accuracy of bed scales compared to standing scales and studies suggest large discrepancies between the two.^5^ Inaccurate weight measurement may lead to medical error, unreliable volume assessment and potentially inappropriate discharge. The utilization of standing scales in lieu of bed scales may improve patient outcomes and overall quality of care in the treatment of ADHF by providing more accurate body weight measurements through a hospitalization. The purpose of this study was to compare and quantify differences in standing and bed scales in patients hospitalized for ADHF and its derivatives.

## Methods

### Study Design

A method-comparison design was used to compare bed and standing scale weights. The primary exposure variable was the type of scale used (bed scale vs. standing scale). The outcome variable was the weight obtained from each scale. Each patient served as his or her own control. Study approval was obtained from the University of Southern California Institutional Review Board (IRB) before data collection.

### Sample Selection

Patients included were adults (age ≥18) admitted to Los Angeles County-University of Southern California Medical Center on days of data collection with a primary or secondary diagnosis of ADHF and its derivatives, including congestive heart failure, acute on chronic heart failure, acute systolic, diastolic or combined systolic and diastolic heart failure, and decompensated heart failure. Patients were located on pre-specified inpatient units, including the Coronary Care Unit (CCU) and select step down and cardiac telemetry units where standing scales had been implemented. Eligible patients were identified via electronic medical record by filtering by unit location and selecting patients by diagnosis. Additional inclusion criteria included the ability to demonstrate capacity, medical clearance to mobilize out of bed and physical ability to get out of bed and stand independently.

Assuming a standard deviation of 3.0 kg and employing t-distribution, sample size was calculated to be 44 with a power of 0.8, alpha of 0.05, and an effect size (ES) of 0.6, using the formula [ES = (mean_groupA_ – mean_groupB_) / SD] (see Appendix A). These parameters were based on data from prior studies and expert opinion that an average difference (bias) of greater than ± 0.6 kg and a SD (precision) greater than ± 3.0 kg between the in-bed and standing scale weights were considered clinically significant and large enough to lead to errors in clinical decision-making related to fluid management in this patient population.^6^

### Data Collection

Collection occurred between March and April of 2023. Eligible participants, identified via electronic medical record, were screened for study inclusion criteria, and written informed consent and HIPAA authorization forms were obtained. Certified interpreters assisted in translation for communication with non-English-speaking patients.

Standard-of-care at our institution requires nursing staff to calibrate bed scales with a standard amount of linen (one pillow, sheet, and blanket) prior to weighing new patients. Research investigators did not repeat the calibration process. Study participants were first weighed on the bed scale (Progressa model, Hill-Rom, Chicago, IL) with the standardized amount of bed linen. Additional items on the bed such as extra linens, patient belongings, and medical equipment were removed to ensure accuracy. Standing weights were then obtained utilizing bedside standing scales (seca 876, Chino, CA) with study team assistance when indicated. We aimed to compare 44 sets of measurements. Data sets were summed together between all participants.

### Analysis

Each set of bed scale and standing scale weights were compared. Standard descriptive analytics were performed. The mean weight difference (bias), precision (SD) and limits of agreement (±2 SD) were calculated and graphed using the Bland-Altman method.

## Results

43 patients participated in the study between March and April of 2023 and a total of 51 pairs of bed scale and standing scale weights were obtained. Eight patients were weighed more than once, as they remained admitted on subsequent days of data collection. By increasing the sample size to 51, our power increased to 0.86. Standing weight values ranged from 43.4 to 117 kg, with a mean of 74.0 ± 32.2 kg (95% CI, 41.8 – 106). Weight differences are detailed in Table I and displayed on a Bland-Altman graph in Figure 1. The bias ± precision between weights was 1.42 ± 1.18 kg (95% CI, -0.894 – 3.73), exceeding the recommended bias value. In addition, 71% (n=36) of participants’ weights differed by more than 0.6 kg between the bed and standing scale, 22% (n=11) differed by more than 2.0 kg, and 5.9% (n=3) differed by more than 4.0 kg (see Table I).

**Table I.** Weights for Study Participants.

|  | **Mean ± SD**<br>**Weights (kg)** | **Difference**<br>**Between In-**<br>**Bed and**<br>**Standing**<br>**Weights**<br>**(bias ±**<br>**precision)**<br>**(kg)** | **Number of**<br>**Weight**<br>**Differences $>$**<br>**$\pm 0.6$ kg** | **Number of**<br>**Weight**<br>**Differences**<br>**$> \pm 2.0$ kg** | **Number of**<br>**Weight**<br>**Differences ≥**<br>**$\pm 4.0$ kg** |
| --- | --- | --- | --- | --- | --- |
| **Standing**<br>**Scale** | 74.0 ± 32.2 | | | | |
| **Bed Weight** | 74.4 ± 32.5 | 1.42 ± 1.18 | 36 (71%) | 11 (22%) | 3 (5.9%) |

**Figure 1.**
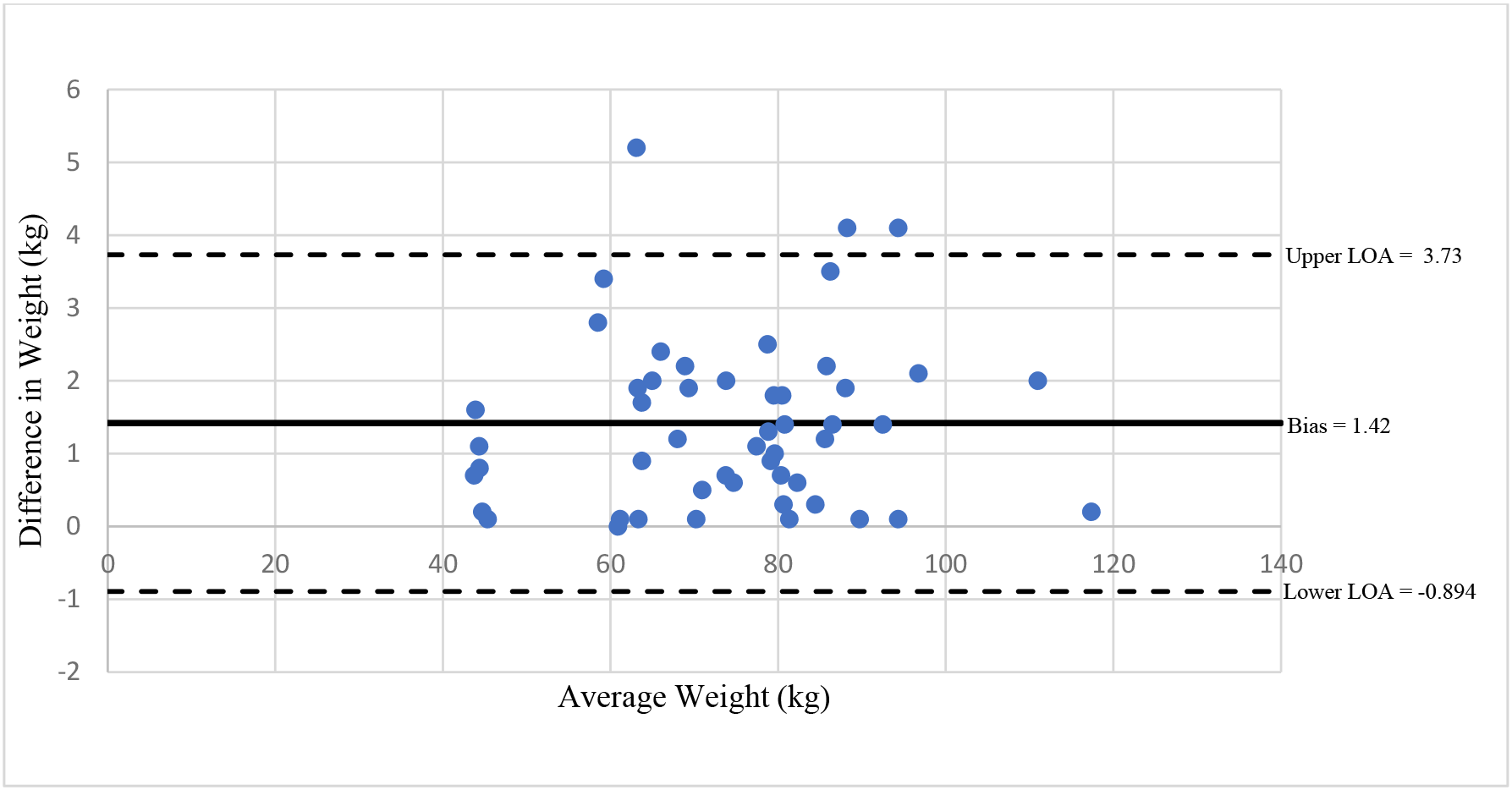
Bland-Altman Plot for Comparison of Bed and Standing Scale Weights. X-axis: average weight of each bed and standing scale weight pair. Y-axis: weight difference between each bed and standing scale weight pair. LOA = limit of agreement.

## Discussion

Weight measurement is critical in the treatment of patients admitted with ADHF. While bed scale weights were precise, they produced inaccurate weights compared to standing scale weights in our study. It is worth noting that accuracy is perhaps more important than precision in this context, as patients are often transferred to different rooms with different scales over the course of admission and medications are dosed based on these recorded weights. There are a multitude of factors that can contribute to user error when weighing patients on bed scales, such as inadequate bed calibration prior to patient admission and lack of attention to having only the standard amount of linen, clothing, and equipment in place during in-bed weight measurement. A study by Schneider et al. comparing electronic bed weighing versus daily fluid balance changes highlighted the contribution of items left in the bed on weight accuracy.^7^ For patients with bed linens and/or medical equipment that could not be removed from the bed due to patient safety and/or comfort, the extra items averaged 3.5 ± 1.4 kg for each patient.^7^ Findings from this study suggest that beds may not be properly calibrated prior to weighing when extra items are left on the bed, leading to inaccurate weights.

Inaccurate weights obtained during admission can have profound medical implications. For example, weight-based medications such as anticoagulants and insulin can cause significant harm when dosed incorrectly. Additionally, the discharge weight of patients hospitalized with ADHF is often used as a reference point for outpatient volume monitoring, as clinical guidelines recommend such patients to be euvolemic at discharge. As even a weight gain of 2 pounds can be associated with decompensation and necessary readmission, an inaccurate discharge weight may interfere with outpatient volume monitoring recommendations.^8^

Further opportunities exist for improved standardized processes of bed calibration, in-bed weighing, and nursing staff education for obtaining accurate weights. When patients are able to ambulate independently, standing scales should be used to provide more accurate weight measurement and estimation of body volume status.

### Limitations

This study may be limited by the particular brand of bed scales present and the specificity to this institution. It may also be limited by research investigators not repeating the bed calibration process. However, this more accurately reflects the clinical reality of how in-bed scales are calibrated in current practice. In the future, this study could be replicated by comparing bed weights before and after calibration to more directly evaluate the effect of accurate calibration. Further, results of this study may not be generalizable to other facilities if the staff follow more standardized weighing procedures.

## Conclusion

Bed scales are inaccurate compared to standing scales when weighing patients admitted and treated for ADHF. When available, standing scales are preferred as bed scale inaccuracies have the potential to lead to errors in medical management.

## Data Availability

The participants of this study did not give written consent for their data to be shared publicly, so supporting data is not available.

## Acknowledgements

None

## Supplemental Material

Table I

Figure 1

Appendix A

### Appendix A

The following assumptions have been made:

Effect Size (ES) of 0.6,

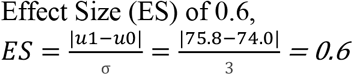

- In-Bed Weight = 75.8 kg, with σ1 = standard deviation 1 = 3, and σ2 = standard deviation 2 = 1.5
- Standard Weight = 74 kg, with σ1 = standard deviation 1 = 3, and σ2 = standard deviation 2 = 1.5

Sample size calculation was based on the following assumptions and the following formula:

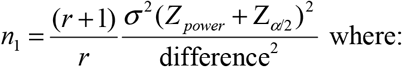

*n*_1_ = size of smaller group

r = ratio of larger group to smaller group

σ = standard deviation of the characteristic

difference = clinically meaningful difference in means of the outcome (1.8)

Z_power_ = corresponds to power (.84 = 80% power)

*Z*_a/2_ = corresponds to two-tailed significance level (1.96 for *α* = .05)

σ1 = standard deviation 1 = 3

σ2 = standard deviation 2 = 1.5

**Table II.**
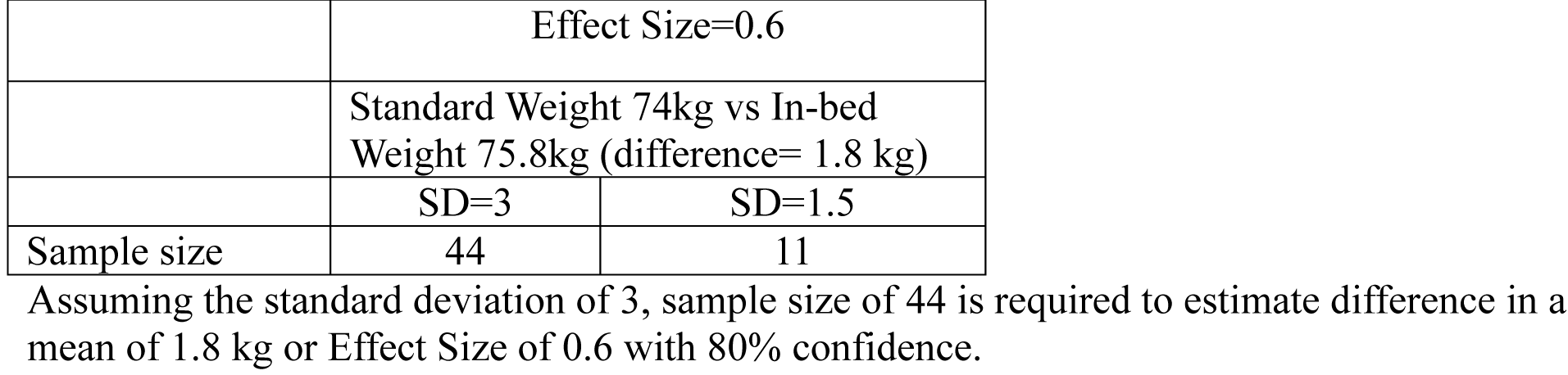
Sample Size table at Power =0.80 and Type I error (*α*) or level of significance=5%.

**Table III.** Power table at Type I error (*α*) or level of significance=5%.

|  | Effect Size=0.6 |  |
| --- | --- | --- |
|  | Standard Weight 74kg vs In-bed Weight 75.8kg (difference= 1.8 kg) |  |
|  | SD=3 | SD=1.5 |
| Sample size of 50 | 85.2% | >99.99% |
| Sample size of 51 | 85.8% | >99.99% |

Current sample size of 51 can detect the effect size of 0.6 with 85.8% confidence.

